# The Association Between IT-Profession-Specific Stressors and Mental Health Conditions Plus the Role of Mental Health Literacy in Help-Seeking

**DOI:** 10.1101/2025.02.24.25322785

**Authors:** Edlin Garcia Colato, Nianjun Liu, Angela Chow, Catherine Sherwood-Laughlin, Jonathan T. Macy

## Abstract

**Background:** The Information Technology (IT) sector is growing and encompasses all professions, from leisure and recreation to hospitals and emergency response groups. IT professionals are experiencing increased threats (e.g., ransomware attacks), but little is known about the relationship between these IT-profession-specific stressors and the professionals’ mental health. This study aimed to 1) estimate the associations between IT-profession-specific stressors and anxiety, depression, and stress, and 2) examine the role of mental health literacy (MHL) as a mediator of the relationship between depression, anxiety, stress, and help-seeking.

**Methods:** Between February and May 2023, 388 IT professionals working in the US were surveyed online. Participants reported demographic characteristics, and their MHL, mental health symptoms, and help-seeking intentions were assessed with the following scales: MHL-in the workplace (MHL-W), Center for Epidemiological Studies Depression-10 (CESD-10), Generalized Anxiety Disorder-7 (GAD-7), Perceived Stress Scale-10 (PSS-10), and the mental help seeking intention scale (MHSIS).

**Results:** Respondents who had experienced ransomware attacks in the past year reported significantly higher symptoms of depression, anxiety, and stress. Adapting to rapid changes in technology and business requirements was associated with higher levels of stress. MHL was found to partially mediate the relationship between depression and help-seeking, but not between anxiety or stress and help-seeking.

**Conclusion:** These findings provide insight into the workplace stressors that pose a greater psychological health risk for IT professionals. These results emphasize the important role of MHL in helping facilitate the connection between depressive symptoms and help-seeking.

## Introduction

Established in the 1950s, Information Technology (IT) is defined as the “use of computer systems or devices to access information” for both business and personal operations, such as “storing, retrieving, accessing or manipulating information” (CompTIA n.d.). The efforts of IT workers to maintain business devices can sometimes involve high-stress exposures such as viewing illicit content and mitigating ransomware attacks (Boehm et al. 2022). Technologies have become persistent targets for hackers and cybercriminals who seek vulnerabilities in networks (Samtani et al. 2017). Ensuring the safekeeping of technology means that some IT professionals are in a constant state of high alert. The high levels of stress experienced by IT workers are reflected in the findings from a recent assessment of challenges and stressors in the information security sector. According to the 2022-2023 Chartered Institute of Information Security (CIISec) report, 22% of 302 UK respondents reported working above 48 hours per week, with 8% working more than 55 hours weekly (Chartered Institute of Information Security 2023). Furthermore, 32% reported they were kept awake by the worries of a potential cyber-attack on their organization, up from 22% in 2022 (Chartered Institute of Information Security 2022). Over two-thirds of CIISec survey respondents believe there will be an increase in the frequency and impact of ransomware attacks (Chartered Institute of Information Security 2022).

The existing literature on mental health in IT has largely focused on IT professionals working in Asia (Lim and Teo 1999; Rao and Chandraiah 2012) and Europe (British Interactive Media Association 2019; Chartered Institute of Information Security 2022). Following a ransomware attack, Northwave (2022), a security company based in the Netherlands, conducted a study with 21 of more than 40 of its own Computer Emergency Response Team (CERT) employees and found that mental health can be significantly affected by ransomware attacks. While the Northwave study suggested an elevated risk in mental health issues among IT professionals, another study utilizing the UK Biobank cohort study did not yield similar findings (Lalloo et al. 2022). This first UK-based longitudinal study (2006-2010), which compared the incidences of anxiety and depression between IT and non-IT employees aged 40 and above found that IT professionals had a reduced risk of anxiety and depression compared to their non-IT counterparts (Lalloo et al. 2022). To the best of our knowledge, no such report exists for the IT sector in the US.

Considering the IT professionals who report experiencing symptoms of depression, anxiety, and stress, an important next step is to determine whether these individuals possess a firm knowledge base of mental health and whether they intend to seek help. Mental health literacy (MHL) refers to the knowledge and ability to recognize and identify symptoms related to mental health for preventing mental illness as well as maintaining and promoting mental health (Jorm et al. 1997). Previous research has found a positive correlation between MHL and mental health attitudes (Lee et al. 2020), and young adults with more favorable attitudes towards mental health services are more likely to seek help (Zorrilla et al. 2019).

Past studies have not assessed MHL, mental health attitudes, nor intention to seek help among IT workers in the US. Additionally, previous studies have not considered important factors such as exposure to illicit content, ransomware attacks, hacking, and takedowns which may contribute to the elevated symptoms of depression, anxiety, and stress experienced by IT professionals. A recently published occasional paper from the UK examined first order harms of ransomware on staff and identified the stress reported by the incident responders following the incident (Jamie MacColl et al. 2024). First order harms are “harms to any organization and their staff directly targeted by a ransomware incident” (Chartered Institute of Information Security 2023). There is an urgent need for evidence-based resources and information concerning mental health within the IT workforce community, a domain that remains understudied, especially in the US contexts. Therefore, the present study aimed to 1) test the relationship between IT-profession-specific stressors and anxiety, depression, and stress and 2) to analyze the role of MHL as a mediator for the three selected mental health conditions (anxiety, depression, stress) and help-seeking behaviors.

## Methods

### Participants and Data Collection

Data for this study were collected between February 2023 and May 2023 via an online survey using Qualtrics (University Information Technology Services 2022). Participants were identified via known contacts, SurveyCircle (SurveyCircle 2023), and Prolific (Prolific 2023). The three eligibility criteria for the survey included 1) being at least 18 years old, 2) working in the IT sector in the US, and 3) having at least 12 months of any IT work experience. Electronic written voluntary consent was recorded for all survey respondents at the start of the survey. After respondents completed the 10–15-minute survey, they were given the option to take part in a drawing for one of four $75 Target e-gift cards. To ensure anonymity in the survey responses, respondents were guided to a separate Qualtrics survey where they could provide an email for the random drawing. This study (Protocol #18281) received approval as an exempt study by the researchers’ institution’s Human Subjects and Institutional Review Board. As shown in Figure 1, of the original 483 responses recorded, 23 observations were excluded because of either discontinuing the survey prior to providing consent, being ineligible, or a bot entry. A total of 388 (84.3%) of the remaining 460, who provided consent and were determined to be valid responses, completed the survey.

### Measures

#### Assessment of mental health status

Symptoms of depression were assessed with the 10-item Center for Epidemiologic Studies Depression (CES-D-10) questionnaire (Radloff 1991; Vilagut et al. 2016). The CES-D-10 helps identify individuals at risk of developing clinical depression and has been previously validated against the original full 20-item CESD questionnaire (Shrout and Yager 1989), designed for screening the general population (Perreira et al. 2005; Radloff 1977; Vilagut et al. 2016). The CES-D-10 scores range between 0 and 30; Scores below 10 were recoded to 0 = “no significant symptoms of depression,” and the scores 10 and above were recoded to 1 = “significant symptoms of depression”, creating a binary variable for depression.

Meanwhile, the 7-item generalized anxiety disorder scale (GAD-7) was used to screen for symptoms of anxiety among the respondents (Spitzer et al. 2006). Responses to the seven questions were summed up for a final score ranging from 0-21. For descriptive purposes the following three cut-off points 5, 10, and 15 (0-4 = minimal anxiety, 5-9 = mild anxiety, 10-14 = moderate anxiety, and 15-21 = severe anxiety) were used to show the different categorical levels of severity. However, the optimal cutoff score for screening anxiety via the GAD-7 scale is 10 (Johnson et al. 2019; Spitzer et al. 2006). Therefore, scores 10 and above were coded as 1 = “yes” for anxiety and below 10 were coded as 0 = “no/minimal” for level of anxiety.

Unlike the anxiety and depression scales, the 10-item perceived stress scale (PSS-10) does not translate to having clinical significance; therefore categorizing scores into groups is only for descriptive purposes, and for the analyses the scores were included as a continuous variable (Cohen et al. 1983). Final scores ranged between 0 to 40. For descriptive purposes, scores 0-13 were coded as “low stress”, 14-26 as “moderate stress”, and scores 27-40 as “high stress”. Increased perceived stress is reflected by higher scores.

#### IT-profession-specific Stressors

For research question #1, 12 past-year IT-profession-specific stressors were identified based on the feedback solicited from two IT-professionals with a combined 35 years of IT experience. The list of the 12 stressors curated by the IT-professionals are: 1) ransomware attacks, 2) exposure to illicit content, 3) takedowns, 4) handling sensitive data and cybersecurity threats, 5) making critical technology decisions with limited information, 6) adapting to rapid changes in technology and business requirements, 7) pressure to solve complex technical issues, 8) constant need to stay up to date with technology, 9) dealing with unexpected system failures and outages, 10) dealing with leadership that does not wish to invest in or be inconvenienced by cybersecurity initiatives, 11) balancing security and usability, and 12) working with limited resources (e.g., budget and personnel).

Respondents were asked a single question “Which of the following on-the-job stressors have you experienced in the past year?” and to select all that apply from the list of stressors provided. The 12 stressors were measured individually as binary variables (yes/no) and a separate count variable was created to identify the total number of the 12 stressors experienced by each respondent (scores ranging from 0 to 12).

#### Mental Health Help-seeking Intentions

For research question #2, the primary outcome was mental health help seeking intentions which was measured using the mental help seeking intention scale (MHSIS) (Hammer and Spiker 2018). Final MHSIS scores range between 1 and 7. Higher scores indicate greater intention to seek help from a mental health professional (Hammer and Spiker 2018).

#### Mental Health Literacy in the Workplace

The mental health literacy tool for the workplace (MHL-W) is comprised of four vignettes depicting a hypothetical coworker’s behavior in the workplace; each vignette has four questions measuring the four distinct MHL concepts (Moll et al. 2017). The following four variables were rated on a scale from 1 (very low) to 5 (very high): 1) level of knowledge in being able to recognize a specific disorder – “*What might be happening with [him/her]*”, 2) level of knowledge and beliefs regarding risk factors and prevention –“*How you could prevent the situation from becoming worse*”, 3) level of knowledge and attitudes about help-seeking – “*What you should say or do in the situation*”, and 4) level of knowledge and beliefs regarding interventions – “*Resources or services that might be helpful*”. The values for all 16 questions were summed up for a final MHL-W score, with possible scores ranging from 16 to 80. Higher scores indicate a greater self-reported knowledge of mental health.

### Demographics

Demographic data were collected on the following characteristics: age, race (Black or African American, American Indian or Alaska Native, Asian or Asian American, and Native Hawaiian or Other Pacific Islander, White, and Other), ethnicity, sex at birth (female vs. male), relationship status, education, and income level groups. Data regarding health-related characteristics included questions about health insurance and mental health history. Work-related background information included: IT job role/title, percentage of daily job responsibilities that involved cybersecurity work, average hours worked per week, and geographic region of primary workplace.

### Covariates

Covariates included age, sex, race, ethnicity, education, income, health insurance, and mental health history.

### Data Analysis

Descriptive statistics, mean and standard deviation (SD) for continuous variables, and frequencies and percentages for categorical variables were computed by respondent sex at birth. A two-sided p-value of less than 0.05 is considered significant.

The following models were conducted to address the first hypotheses and identify which IT-profession-specific stressors are associated with anxiety, depression, and stress: 1) unadjusted and adjusted logistic regression models to examine the relationship between each of the 12 IT-profession specific stressors and depression (CESD-10), 2) unadjusted and predictor-adjusted logistic regression models to test for the relationship between each of the 12 IT-profession specific stressors and anxiety (GAD-7), and 3) unadjusted and predictor-adjusted linear regression models to test for the association between each of the 12 IT-specific stressors and stress (PSS-10).

A confirmatory factor analysis (CFA) was conducted for the MHL-W because the workplace questionnaire had previously been used only in a single workplace setting in healthcare. The CFA aimed to test whether each of the four questions per vignette loaded onto the intended corresponding MHL constructs. Results of the CFA are presented in supplemental material.

### Mediation Analysis

Mediation analyses were conducted using the ‘medsem’ package available on Stata. Outputs for the structural equation modeling for the mediation analyses includes a modified version of the Baron and Kenny (1986) approach using the Sobel test for assessing the indirect effects. The output also includes the Zhao et al. (2010) method which uses the Monte Carlo resampling approach for assessing the indirect effects. Descriptive statistics, CFA, mediation and all other analyses were conducted using Stata SE 17.0 (StataCorp 2021).

## Results

The majority of the 388 study self-identified as male (73.2%), were white (77.8%), were of non-Hispanic/Latinx ethnicity (89.2%), were married, in a domestic relationship, or engaged (59.5%), had earned a bachelor’s degree or higher (71.9%), and earned more than $75,000 a year (62.9%).

### Outcome Data

The majority (63.7%) were below the minimum cutoff score of 10 for depression as assessed by the CES-D-10. Similarly, for anxiety 53.3% of the respondents reported having less than mild symptoms of anxiety. The mean PSS-10 score was 15.1 (SD 8.0), representing a moderate level of stress. For mental health history, most (71.9%) of the respondents reported having no known previous mental health diagnosis.

### Main Results

#### IT-profession-specific stressors and Depression

The respondents who reported past year exposure to ransomware or working with leadership that did not wish to invest in nor be inconvenienced by cybersecurity initiatives in the past year were more likely to have significant symptoms of depression, including after controlling for the covariates (Table 1). In addition, respondents who reported making critical technology decisions with limited information in the past year were more likely to report significant depressive symptoms compared to the individuals who did not report having to make critical technology decisions. The total number of stressors experienced in the past year also had a significant positive association with depressive symptoms (odds ratio [OR] = 1.12, 95% confidence interval [CI] = 1.02, 1.23, p-value = 0.01), after controlling for the covariates. None of the remaining IT-profession-specific stressors experienced in the past year were individually associated with significant symptoms of depression.

**Table 1.** Regression results for IT-profession specific stressors and depression, anxiety, and perceived stress.

|  | CESD-10 |  | GAD-7 |  | PSS-10 |  |
| --- | --- | --- | --- | --- | --- | --- |
| | OR (95% CI) | adjusted<sup>a</sup> OR (95% CI) | OR (95% CI) | adjusted<sup>a</sup> OR (95% CI) | $\beta$ (95% CI) | adjusted<sup>a</sup> $\beta$ (95% CI) |
| Ransomware |  |  |  |  |  |  |
| No | Ref | Ref | ref | ref | ref | ref |
| Yes | 2.32 (1.21, 4.42)* | 2.05 (0.88, 4.78) | 1.61 (0.76, 3.39) | 1.53 (0.56, 4.21) | 2.15 (-0.40, 4.70) | 0.93 (-1.82, 3.67) |
| Illicit content |  |  |  |  |  |  |
| No | Ref | Ref | Ref | Ref | Ref | Ref |
| Yes | 3.05 (1.60, 5.81)** | 3.94 (1.58, 9.86)** | 2.77 (1.37, 5.62)** | 6.78 (2.24, 20.50)** | 2.67 (0.17, 5.17)* | 2.98 (0.15, 5.82)* |
| Takedowns |  |  |  |  |  |  |
| No | Ref | Ref | Ref | Ref | ref | Ref |
| Yes | 1.20 (0.42, 3.41) | 0.86 (0.17, 4.23) | 1.99 (0.66, 5.99)* | 4.45 (0.83, 23.97)* | 3.29 (-0.72, 7.31) | 3.72 (-1.12, 8.57) |
| Sensitive data |  |  |  |  |  |  |
| No | Ref | Ref | Ref | Ref | ref | Ref |
| Yes | 1.20 (0.72, 2.00) | 1.33 (0.68, 2.59) | 1.16 (0.63, 2.15) | 1.47 (0.65, 3.35) | 1.21 (-0.73, 3.15) | 2.18 (0.12, 4.23)* |
| Tech decisions |  |  |  |  |  |  |
| No | Ref | Ref | Ref | Ref | Ref | Ref |
| Yes | 1.54 (0.92, 2.60) | 2.93 (1.42, 6.06)** | 1.41 (0.76, 2.63) | 2.11 (0.90, 4.95) | 1.67 (-0.28, 3.61) | 2.95 (0.92, 4.99)** |
| Rapid changes |  |  |  |  |  |  |
| No | Ref | Ref | Ref | Ref | Ref | ref |
| Yes | 1.44 (0.84, 2.49) | 2.08 (0.99, 4.34) | 1.16 (0.61, 2.22) | 1.24 (0.52, 2.93) | 2.19 (0.17, 4.20)* | 2.85 (0.73, 4.98)** |
| Tech issues |  |  |  |  |  |  |
| No | Ref | Ref | Ref | Ref | ref | Ref |
| Yes | 1.11 (0.62, 1.99) | 2.23 (0.98, 5.07) | 0.92 (0.46, 1.84) | 1.61 (0.61, 4.27) | -0.12 (-2.36, 2.12) | 1.22 (-1.20, 3.65) |
| Staying up to date |  |  |  |  |  |  |
| No | Ref | Ref | Ref | ref | Ref | Ref |
| Yes | 0.79 (0.46, 1.35) | 0.85 (0.42, 1.75) | 0.72 (0.38, 1.34) | 0.72 (0.29, 1.75) | -0.53 (-2.60, 1.53) | -0.02 (-2.25, 2.20) |
| System failures |  |  |  |  |  |  |
| No | Ref | Ref | Ref | Ref | Ref | Ref |
| Yes | 0.96 (0.56, 1.67) | 0.92 (0.46, 1.86) | 0.90 (0.47, 1.73) | 1.04 (0.45, 2.43) | 1.36 (-0.72, 3.44) | 1.09 (-1.07, 3.25) |
| Leadership issues |  |  |  |  |  |  |
| No | Ref | Ref | Ref | ref | Ref | Ref |
| Yes | 1.38 (0.78, 2.42) | 2.34 (1.06, 5.17)* | 1.72 (0.89, 3.31) | 3.05 (1.16, 8.00)* | 0.65 (-1.54, 2.85) | 2.08 (-0.31, 4.48) |
| Balance |  |  |  |  |  |  |
| No | ref | Ref | Ref | ref | Ref | ref |
| Yes | 0.89 (0.54, 1.49) | 1.26 (0.65, 2.46) | 0.82 (0.44, 1.51) | 1.16 (0.50, 2.68) | 0.61 (-1.33, 2.56) | 2.72 (0.67, 4.77)** |
| Limited Resources |  |  |  |  |  |  |
| No | Ref | Ref | Ref | Ref | Ref | Ref |
| Yes | 1.18 (0.69, 2.00) | 1.56 (0.80, 3.04) | 1.13 (0.60, 2.14) | 1.33 (0.60, 2.98) | 2.06 (0.07, 4.04)* | 2.39 (0.36, 4.41)* |
| Total Stressors | 1.08 (0.98, 1.19) | 1.18 (1.04, 1.35)** | 1.06 (0.95, 1.18) | 1.16 (1.00, 1.35) | 0.40 (0.05, 0.76)* | 0.67 (0.29, 1.05)** |

<sup>a</sup>Adjusted for age, sex, race, ethnicity, education, income, health insurance, and mental health history \*p<0.05; \*\*p<0.01; \*\*\*p<0.001

#### IT-profession-specific Stressors and Anxiety

Past year exposure to takedowns as well as balancing security and usability remained positively associated with symptoms of anxiety after adjusting for the covariates. Past year exposure of dealing with leadership that does not wish to invest in or be inconvenienced by cybersecurity issues were more likely to report moderate symptoms of anxiety compared to those who did not have problems with leadership-cybersecurity issues, after controlling for covariates.

#### IT-profession-specific Stressors and Stress

Past year exposure to ransomware, takedowns, adapting to rapid changes in technology and business requirements, and increased number of stressors were all still positively associated with higher levels of stress after adjusting for the covariates. After controlling for the covariates, past year experience of making critical technology decisions with limited information was significantly associated with increased levels of stress compared to those who did not experience making critical technology decisions in the past year.

### Mediation Analyses

Mediation analysis was performed to assess the mediating role of MHL-W in the relationship between depression, anxiety, and stress scores (separately) and help-seeking intentions (MHSIS). The results (shown in Table 2) revealed a significant indirect effect between depression and MHSIS (/3 = - 0.105, z = −1.99, p = 0.047). The total effect of depression on MHSIS was significant (/3 = −0.794, z = - 4.14, p <0.001), with the inclusion of the mediator (MHL-W), the effect of depression on MHSIS was still significant (/3 = −0.688, z = −3.69, p < 0.001). Per the Baron and Kenny (1986) and the Zhao et al. (2010) approaches, MHL-W partially mediated the relationship between depression and help-seeking intentions.

**Table 2.** Mediation results for MHL-W and depression, anxiety, and stress.

| Mediation Analysis for CESD-10 → MHL-W → MHSIS |  |  |  |  |  |  |  |  |  |  |  |
| --- | --- | --- | --- | --- | --- | --- | --- | --- | --- | --- | --- |
| Total effects (CESD-10 → MHSIS) |  |  | Direct effects (CESD-10 → MHSIS) |  |  | Indirect effects of CESD-10 on MHSIS |  |  |  |  |  |
| coefficient | z-value | p-value | coefficient | z-value | p-value | coefficient | SE | z value | P value | Percentile bootstrap 95% CI |  |
|  |  |  |  |  |  |  |  |  |  | Lower | Upper |
| -0.794 | -4.14 | <0.001 | -0.688 | -3.69 | <0.001 | -0.105 | 0.053 | -1.985 | 0.047 | -0.209 | -0.001 |

| Mediation Analysis for GAD-7 → MHL-W → MHSIS |  |  |  |  |  |  |  |  |  |  |  |
| --- | --- | --- | --- | --- | --- | --- | --- | --- | --- | --- | --- |
| Total effects (GAD-7 → MHSIS) |  |  | Direct effects (GAD-7 → MHSIS) |  |  | Indirect effects of GAD-7 on MHSIS |  |  |  |  |  |
| coefficient | z-value | p-value | coefficient | z-value | p-value | coefficient | SE | z value | P value | Percentile bootstrap 95% CI |  |
|  |  |  |  |  |  |  |  |  |  | Lower | Upper |
| -0.069 | -3.83 | <0.001 | -0.070 | -4.06 | <0.001 | 0.001 | 0.005 | 0.20 | 0.842 | -0.009 | 0.011 |

| Mediation Analysis for PSS-10 → MHL-W → MHSIS |  |  |  |  |  |  |  |  |  |  |  |
| --- | --- | --- | --- | --- | --- | --- | --- | --- | --- | --- | --- |
| Total effects (PSS-10→ MHSIS) |  |  | Direct effects (PSS-10→ MHSIS) |  |  | Indirect effects of PSS-10 on MHSIS |  |  |  |  |  |
| coefficient | z-value | p-value | coefficient | z-value | p-value | coefficient | SE | z value | P value | Percentile bootstrap 95% CI |  |
|  |  |  |  |  |  |  |  |  |  | Lower | Upper |
| -0.048 | -4.23 | <0.001 | -0.044 | -3.94 | <0.001 | -0.005 | 0.003 | -1.51 | 0.130 | -0.011 | 0.001 |

On the contrary, the results for mediation analyses for anxiety and stress did not support MHL-W as a mediator for anxiety or stress and help-seeking intentions. Although there was a significant relationship between MHL-W and MHSIS, there was no significant relationship between GAD-7 nor PSS-10 scores and MHL-W.

## Discussion

### Key Results

This study was conducted to 1) test which IT-profession-specific stressors were associated with depression, anxiety, and stress, and 2) examine the impact of depression, anxiety, and stress on help-seeking as mediated by mental health literacy. For the first objective, testing which IT-profession-specific stressors were associated with anxiety, depression, and stress, we found that IT professionals with past year exposure to ransomware attacks were more likely to have significant symptoms of depression, anxiety, and stress. This finding aligns with Northwave’s report on the impact of a ransomware attack had on 21 of their employees, in which significant stress was experienced immediately after the incident and throughout the following year (Northwave 2022). This adds to the ransomware literature, which is heavily focused on the financial impacts of individuals and organizations and the psychosocial costs of the victims, rather than considering the individuals working tirelessly to mitigate the problem (Chen 2022; van Boven et al. 2023).

The pressure to solve complex technical issues, the constant need to stay up to date with technology, and dealing with unexpected system failures and outages were not associated with depression, anxiety, or stress. This is likely because these stressors, except for the unexpected system failures and outages, are typical daily job activities and expected tasks and responsibilities for individuals working in the IT sector. Staying up to date with technology and solving complex technical issues are part of the allure of working in technology. These may act as positive stressors, potentially increasing stress while also promoting professional and personal growth, rather than predominantly negative experiences for IT professionals (Jensen 2019). The influence of positive and welcomed anticipated stressors among IT professionals requires further exploration.

Interestingly, exposure to illicit content was not associated with depression, anxiety, or stress in this study. Previous literature has found that exposure to illicit content (e.g., exploitive, violent, and/or abusive content) can have psychologically detrimental effects on children and adolescents (Meates 2020). While similar associations have not been extensively documented among adults, Federal Bureau of Investigation (FBI) officers have developed coping strategies to deal with exposures to illicit content. For FBI officers who are employed in this line of work, the skill of compartmentalization is an essential key for their capacity to sustain their professional roles (Spotlights 2011). Future research should look into the type and amount of illicit content exposures to increase our understanding of when and how this content becomes harmful, specifically among adults and as an occupational hazard.

Regarding the second objective, aiming to assess to what extent MHL mediated the relationship between anxiety, depression, or stress and help-seeking behaviors, mental health literacy in the workplace only partially mediated the relationship between depression and help-seeking intentions, but not between anxiety or stress and help-seeking intentions. Unlike depression and anxiety, stress is not a medical diagnosis, but rather an automatic bodily response that is experienced after certain events. Therefore, it often does not require seeking medical help. Stress is experienced daily and stress levels constantly fluctuate (World Health Organization 2023). Chronic stress, on the other hand, can increase the risk for depression and anxiety (Tafet and Bernardini 2003), but the PSS-10 instrument used in this study captured past-week perceived stress, not chronic stress.

Compared to the 2019 UK industry report which indicated 28% of survey respondents experienced past year anxiety and depression (British Interactive Media Association 2019), the sample in the current study shared a similar percentage, with 28.1% reporting ever having been diagnosed with a mental health condition. On the other hand, for current depression and anxiety via the CESD-10 and GAD-7 scores, 29.3% of respondents were assessed as having recent symptoms of clinically significant depression and mild and above symptoms of anxiety. These numbers are much lower than the 81% reported by BIMA. Differences in these numbers could be due to how the questions were formulated. This study uses two validated instruments to capture symptoms, while the UK industry survey asked respondents to self-report past year (12-months) anxiety and depression experiences.

Unfortunately, the results of this study are not directly comparable to those of the UK Biobank cohort study. It is important to note that the UK Biobank cohort study, in addition to being a longitudinal study with a UK sample, measured depression and anxiety with a single assessment tool, Patient Health Questionnaire (PHQ)-4, while this US focused study included the CESD-10 and GAD-7. Nevertheless, the current study was the first to assess the relationship between IT-profession-specific stressors and depression, anxiety, and stress, aiming to shed light on their associations. It also attempted to uncover the mediating role of mental health literacy between mental health conditions and help-seeking intentions.

### Limitations and Generalizability

There are a few limitations that must be noted. First, this is a cross-sectional study design, and although a mediation analysis was conducted, direction nor cause and effect of the relationships can be determined. The relationship found are associations and must not be regarded as causal. Furthermore, the instrument, MHL-W, used to ascertain MHL is self-reported level of knowledge, which might be over-inflated.

Although the sample resembled the demographic characteristics of the IT professionals by sex (73.2% male vs. 26.8% female) (White 2023), this sample is slightly overrepresented by White respondents (77.8%). The 2019 US Census Bureau estimates high technology industries are comprised of primarily White (68.5%), followed by 14% Asian American IT professionals (U.S. Equal Employment Opportunity Commission n.d.). Although this sample is not representative of the demographic characterization of the current IT sector, the goal of this study was to understand the relationship between the exposures and outcome regardless of demographic information.

Despite the limitations, the results of this exploratory study are promising in that they provide insight to the types of stressors that pose a greater psychological health risk for the IT professionals. Again, the possible high-risk stressors identified in this study include past year exposure to ransomware attacks, making critical technical decisions with limited information, and dealing with leadership that is not interested in cybersecurity initiatives. These results might also be beneficial to IT leaders when considering what response plans are in place to help mitigate the psychosocial impacts these exposures may have on their IT professionals. As been shown previously, leadership training in mental health resources impacts the employee’s willingness and actual use of those resources (Dimoff and Kelloway 2019). Furthermore, the findings highlight the important role of MHL in helping facilitate that connection between experiencing significant symptoms of depression and seeking help to address the depressive symptoms.

## Conclusion

As the IT workforce continues to expand throughout many other sectors, thus increasing IT professional opportunities for employment, the lack of comprehensive mental health support and resources in some sectors could have negative consequences for their overall quality of life. Their health and well-being are crucial not only from a worker and an industry-level perspective, but also because of the significance of their job roles. Gaining insight into their mental health needs can facilitate the development of strategies for implementing programs aimed at improving and/or maintaining good mental health.

Before leaders can identify solutions and resources for their team, we need leaders to be on board to support their team. Given the importance of employee safety, health, and well-being, results from this study can aid IT leaders in identifying situations where IT professionals could be at an elevated risk due to workplace stressors. This awareness would enable them to proactively provide resources and support in a timely fashion. For IT-specific stressors, it is not a matter of if, but rather when these situations such as a ransomware attack will occur. Therefore, having a plan that not only focuses on the attack itself, but the human being the computer will be essential for promoting health among IT professionals.

## Funding

The authors received no external financial support for the research, authorship, and/or publication of this article.

## Conflict of Interest

The authors declare no conflict of interest relating to the material presented in this Article. Its contents, including any opinions and/or conclusions expressed, are solely those of the authors.

## Data Availability

The datasets generated during and/or analyzed during the current study are available from the corresponding author upon reasonable request.

## Notes

We have no conflicts of interests to disclose.

### Competing Interest Statement

The authors have declared no competing interest.

### Funding Statement

This study did not receive any funding.

### Author Declarations

Human Subjects and Institutional Review Board of Indiana University Bloomington gave ethical approval for this work (Protocol #18281).

